# Modelling patterns of SARS-CoV-2 circulation in the Netherlands, August 2020-February 2022, revealed by a nationwide sewage surveillance program

**DOI:** 10.1101/2022.05.25.22275569

**Authors:** Michiel van Boven, Wouter A. Hetebrij, Arno M. Swart, Erwin Nagelkerke, Rudolf F.H.J. van der Beek, Sjors Stouten, Rudolf T. Hoogeveen, Fuminari Miura, Astrid Kloosterman, Anne-Merel R. van der Drift, Anne Welling, Willemijn J. Lodder, Ana M. de Roda Husman

**Affiliations:** Centre for Infectious Disease Control, National Institute for Public Health and the Environment (RIVM), Bilthoven, the Netherlands; Julius Center for Health Sciences and Primary Care, University Medical Center Utrecht, Utrecht, the Netherlands; Center for Marine Environmental Studies (CMES), Ehime University, Ehime, Japan; Institute for Risk Assessment Science (IRAS), Utrecht University, Utrecht, the Netherlands

## Abstract

**Background:** Surveillance of SARS-CoV-2 in wastewater offers an unbiased and near real-time tool to track circulation of SARS-CoV-2 at a local scale, next to other epidemic indicators such as hospital admissions and test data. However, individual measurements of SARS-CoV-2 in sewage are noisy, inherently variable, and can be left-censored.

**Aim:** We aimed to infer latent virus loads in a comprehensive sewage surveillance program that includes all sewage treatment plants (STPs) in the Netherlands and covers 99.6% of the Dutch population.

**Methods:** A multilevel Bayesian penalized spline model was developed and applied to estimate time- and STP-specific virus loads based on water flow adjusted SARS-CoV-2 qRT-PCR data from 1-4 sewage samples per week for each of the >300 STPs.

**Results:** The model provided an adequate fit to the data and captured the epidemic upsurges and downturns in the Netherlands, despite substantial day-to-day measurement variation. Estimated STP virus loads varied by more than two orders of magnitude, from approximately 10^12^ (virus particles per 100,000 persons per day) in the epidemic trough in August 2020 to almost 10^15^ in many STPs in January 2022. Epidemics at the local levels were slightly shifted between STPs and municipalities, which resulted in less pronounced peaks and troughs at the national level.

**Conclusion:** Although substantial day-to-day variation is observed in virus load measurements, wastewater-based surveillance of SARS-CoV-2 can track long-term epidemic progression at a local scale in near real-time, especially at high sampling frequency.

## INTRODUCTION

The SARS-CoV-2 pandemic has posed an unprecedented threat to public health in modern times. The virus can cause the respiratory and systemic disease COVID-19, but in general severity and progression of the disease are variable [1], depending on host risk factors [2] and pathogen variants [3]. It is known that transmission can also occur via asymptomatic or pre-symptomatic individuals [4]. These characteristics hamper effective epidemiological surveillance for the infection dynamics, as case notifications are biased, depending on strain-dependent severity of disease, willingness to get tested, testing capacity, and public health policies. Epidemiological surveillance based on hospital admissions is less biased, but it does not provide an accurate picture in the early and late stages of an epidemic when hospitalisations are rare. This is especially true at a local scale (e.g., municipalities) and for variants causing relatively mild disease (such as the Omicron variant relative to the Delta variant). Serological surveillance for SARS-CoV-2 can provide an unbiased population-level estimate of the fraction of the population that has been infected [5]. Serological surveys, however, are costly and each survey only provides a cross-sectional snapshot of the population at a single time point.

In contrast with earlier applications of wastewater-based surveillance such as the identification of emerging enteroviruses [6], monitoring of SARS-CoV-2 in sewerage can supply quantitative information at a local scale [7]. In the COVID-19 pandemic, the detection of SARS-CoV-2 in feces [8] has accelerated the introduction of wastewater surveillance in a variety of settings, including airports, hospitals, and cities. Wastewater-based surveillance has now been implemented in many countries globally [9]. However, many of these activities are carried out by local governmental bodies or independent research groups and are often restricted to specific locations. This makes it difficult to compare variations over time and geographically in a broader perspective.

Here we report data and analyses from a national wastewater-based surveillance program in the Netherlands. Since September 7^th^, 2020, all sewage treatment plants (STPs) in the Netherlands provide 1 to 4 samples per week to the Dutch National Institute for Public Health and the Environment. Since almost every Dutch household is connected to a sewer, coverage of the program is close to 100%. Samples are subjected to a standardised extraction protocol and analysed within 2-4 days after sampling by real-time RT-PCR. As individual measurements of SARS-CoV-2 RNA in sewage are noisy, inherently variable, and can be left-censored (i.e., not able to detect RNA at low virus loads), our aim is to integrate the available data to provide estimates of the latent true SARS-CoV-2 virus loads over time for all STPs. We provide an integrated analysis of the data from August 1, 2020 (when 60% of the population had already been included) up to and including February 8, 2022 (when more than 99% of the population had been included).

Hence, the time series data cover the winter epidemic of 2020-2021, the 2021 summer surge after many restrictions had been lifted, and the 2021-2022 winter epidemic. Our analyses employ a multilevel Bayesian model with flexible smoothing functions (splines) describing virus load trends at the level of STPs, enabling information sharing both within and between STP time series. We discuss how sewage surveillance could help anticipate and prepare for future epidemics of SARS-CoV-2.

## MATERIALS AND METHODS

### Sampling, RNA extraction, and qRT-PCR

In the Netherlands, sewage of more than 99% of the population is treated at 317 STPs, of which 4 closed between start and end of the period of analysis (Figure 1). Coverage of the program is over 99% of the Dutch population (>17 million) as only a small fraction of the population in the Netherlands is not connected to a sewage system. STPs vary in size from covering approximately 1,000 to over 800,000 inhabitants. Sewage samples are collected using a 24h flow proportional sampling device from which 500-1000 mL is sent to the laboratory. Quantification of SARS-CoV-2 RNA in the samples was based on a standard curve. Details of sampling and experimental procedures will be provided elsewhere (Lodder et al, in preparation).

**Figure 1.**
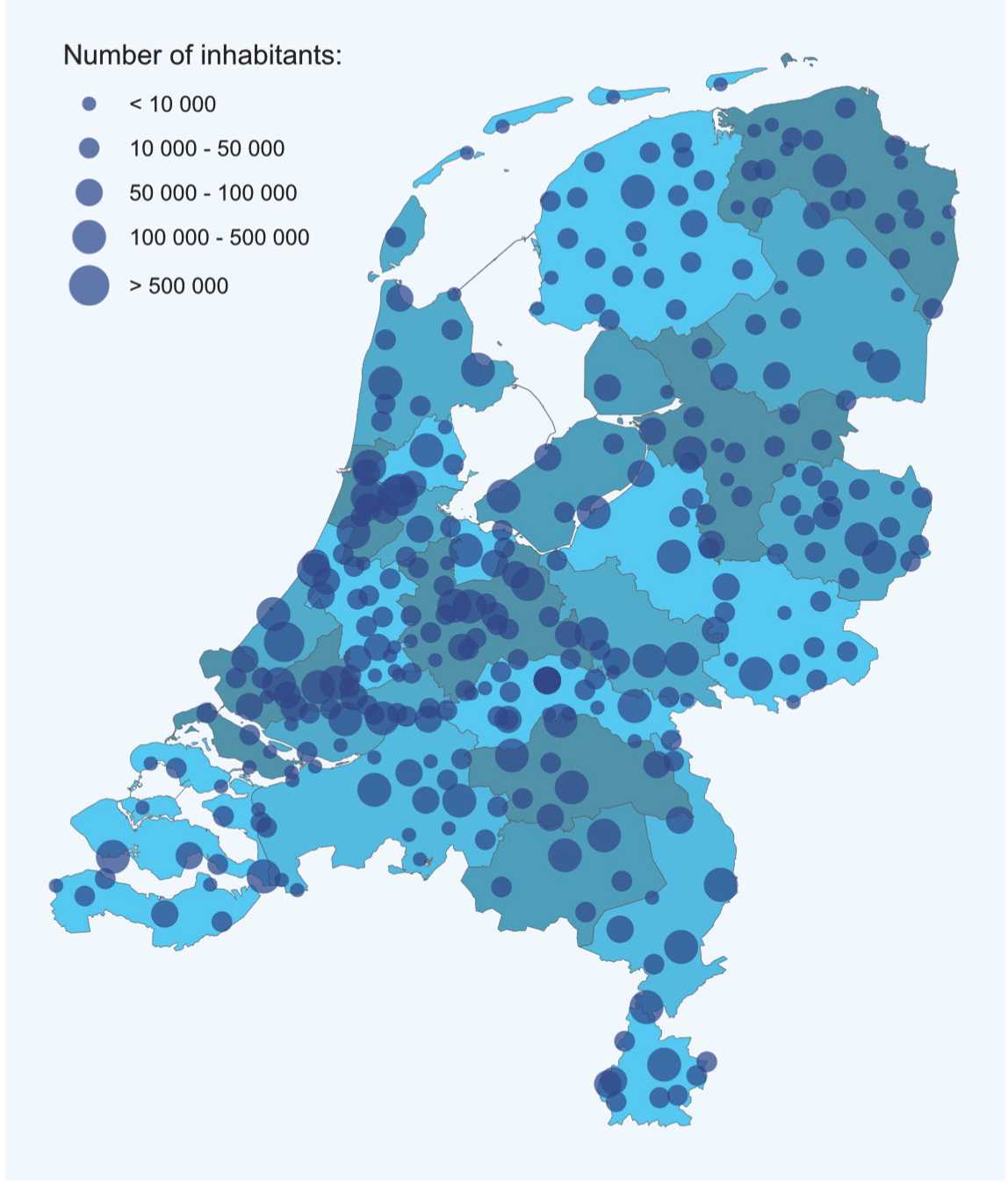
Sewage treatment plants in the Netherlands. Shown are locations of all 313 STPs in 2022 (dots). Sizes of the dots represent the number of persons serviced by the STPs. Background colors represent the 25 safety regions of the Netherlands.

### Statistical analysis

We analyse log_10_-transformed virus concentration data, normalised by 24h sewage water flow rates (unit: virus particles per 100,000 persons per day; henceforth called virus load) [10] using a Bayesian multilevel spline model with random effects at the level of the STPs [11-13]. In the model, the expected log_10_-transformed virus load at time *t* and STP *i, c*_*i*_(*t*), is given by

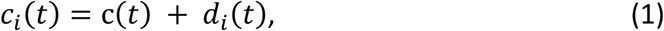

where *c*(*t*) represents the national trend, and *d*_*i*_(*t*) represent the STP-specific deviations. The national trend is modelled with a penalised spline (p-spline) with cubic b-spline basis functions and 15 equidistant knots (yielding 17 regression coefficients). Results presented here are obtained with a first-order random walk (RW1) prior for the regression coefficients [12], using a *N*(12,1) prior distribution for the first regression coefficient and Inverse-Gamma(1, 0.0005) prior distribution for the variance parameter [14]. In this manner, the national log_10_-transformed virus load at the first time point is normally distributed a priori with a mean of 12 (per 100,000 persons per day) and approximate 95% prior range of 10-14 (per 100,000 persons per day). This provides good coverage for virus loads at the first time points (August 2020). In a similar manner, STP-specific deviations from the trend are modelled with cubic b-splines with 15 equidistant knots (yielding 17 coefficients per STP), and with zero-mean normal prior distributions (*N*(0,1)). Hence, plant-specific deviations are a priori expected to be no more than approximately 100-fold (i.e., 2 standard deviations) lower or higher than the trend. We also explored a suite of alternatives, e.g., estimating the variance of the deviations, or replacing the p-spline for the national virus load with a b-spline using *N*(13,1) prior distributions for the regression coefficient. Throughout, we assume that the log_10_-transformed virus load measurements *Y*_*it*_ at plant *i* and time *t* are normally distributed (*Y*_*it*_∼*N*(*c*_*i*_(*t*), *σ*)), and we use an uninformative (improper uniform) prior distribution for *σ* > 0.

The probability of virus detection in a sample is determined by the detection limit of the qRT-PCR, the daily flow rates of water through the sewerage, and possibly also by the composition of the sewage. Hence, there is no fixed predefined cut-off for RNA detection that can be applied to all STPs at all time points, and we include a two-parameter logistic function that determines the probability of detection in the analyses. The parameters *c*_0_ (load at which detection occurs with a probability of 0.5) and *k* (steepness of the detection curve) are estimated. Hence, denoting the probability of detection by 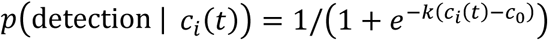, a non-detect adds a contribution 1 − *p*(detection | *c*_*i*_(*t*)) to the likelihood. In this manner, samples in which no RNA is detected tend to pull the estimated load *c*_*i*_(*t*) to lower values. The parameters *c*_0_ and *k* are given uninformative (improper) uniform prior distributions (*c*_0_ > 0 and *k* > 2). The event of a detection is Bernoulli distributed,

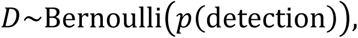

where the dependence of *p*(detection) on the latent estimated true load *c*_*i*_(*t*) is suppressed. Putting it together, the log-likelihood contributions are given by

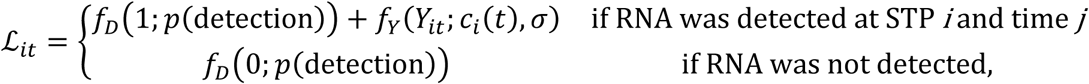

where *f*_*D*_ and *f*_*Y*_ denote the log Bernoulli and log normal probability densities, respectively.

To tie the results at the STP level to other organisational levels (municipality, safety region, national) we apply posterior weighting of the estimated virus loads (i.e., the exponentiated estimated log-loads), where weighting is proportional to the numbers of persons contributing to the various units of organisation. Demographic data is obtained from Statistics Netherlands using the census of 1 January 2020 [15], with the exception of the municipal mergers of January 2021 and 2022 which were manually added to the demographic data. Of note, the analyses reported here include all but the three overseas extraordinary municipalities.

We analyse sewage data of more than 47,000 measurements spanning the period 1 August 2020 (when the majority of STPs had been included in the national program) up to and including 8 February 2022. All analyses are performed with Stan (version 2.21.0) and R (version 4.1.3), using RStan (version 2.21.2) as interface to Stan [16]. We run 10 MCMC chains in parallel and base the analyses on 1,000 samples from well-mixed chains, where we have applied 1/4 thinning to minimise correlations between samples. Scripts, figures, and results underlying figures are available in the online repository, available at https://github.com/rivm-syso/SARS-CoV-2_sewage.

## RESULTS

The Dutch sewage surveillance program was initiated in the first months of 2020. Since its inception, the number of included STPs has increased over time such that 80 STPs provided samples in August 2020, that all 313-317 STPs provided samples from September 2020 onwards, and that the frequency of sampling increased from 1 to 4 samples per week. An overview of the location and size of STPs in the Netherlands is provided in Figure 1, and shows that STPs in the densely populated western parts of the Netherlands are generally significantly larger than those in other regions. Noteworthy, STPs often process sewage from multiple municipalities, and it is not uncommon that a municipality is being served by multiple STPs. Hence, STPs do not follow the hierarchical organisational structure of the Dutch administrative divisions, which makes averaging from STP-level data and results to municipalities, provinces, and safety regions not straightforward.

Figure 2 shows the data (dots) together with the posterior medians (lines) and 95% posterior credible intervals (grey bands) of the latent virus loads for the nine largest STPs. Results for all 317 STPs are available in the online repository at https://github.com/rivm-syso/SARS-CoV-2_sewage. In general, the model describes the data well, and main statistical indicators suggest that the estimation procedure yielded satisfying results (scale reduction factor Rhat close to 1, no divergent transitions, components of the posterior distribution are located well within the support of the prior distributions, strong posterior contraction, no systematic deviations of residuals from the posterior median). Estimated virus loads were generally low in August-September 2020 and are estimated between 10^12^ to 10^13^ virus particles per 100,000 inhabitants per day. At these virus loads a sizeable fraction of samples can yield a non-detect, especially at the lower range of 10^12^. This is illustrated in Figure 3, which shows the probability of detection as a function of virus load. The estimated probability of detection is close to 0 at a virus load of 10^11^, close to 50% at a virus load of 10^12^, and almost 1 at a virus load of 10^13^. Estimated parameters of the detection function are *c*_0_ = 12.1 (95%CrI: 12.1-12.2) for the posterior median of the logistic midpoint, and *k* = 4.2 (95%CrI: 4.0-4.4) for the posterior median of the logistic growth rate.

**Figure 2.**
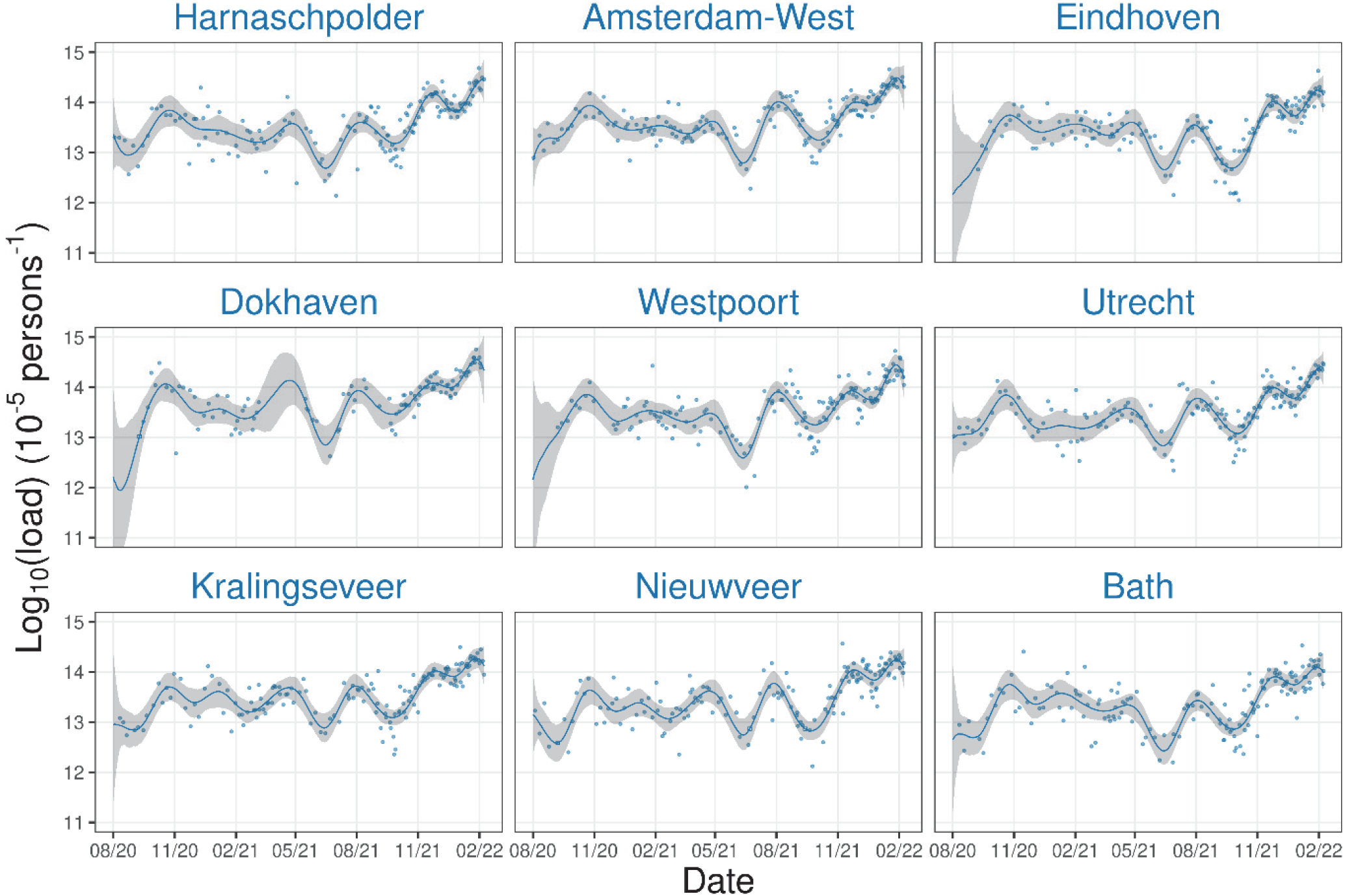
SARS-CoV-2 loads in sewage (dots) and fit of the model (lines) for the nine largest STPs in the Netherlands. STPs are sorted by size (top left to bottom right). Total number of persons connected to these plants is 3,641,251 (Statistics Netherlands, 2020), representing 21.0% of the population covered by all 317 STPs in The Netherlands. Lines show the posterior medians of the fitted virus loads with 95% credible intervals (grey bands). Open dots show samples in which no RNA was detected and are plotted on top of the posterior median. Figures and underlying data for all STPs are available at https://github.com/rivm-syso/SARS-CoV-2_sewage.

**Figure 3.**
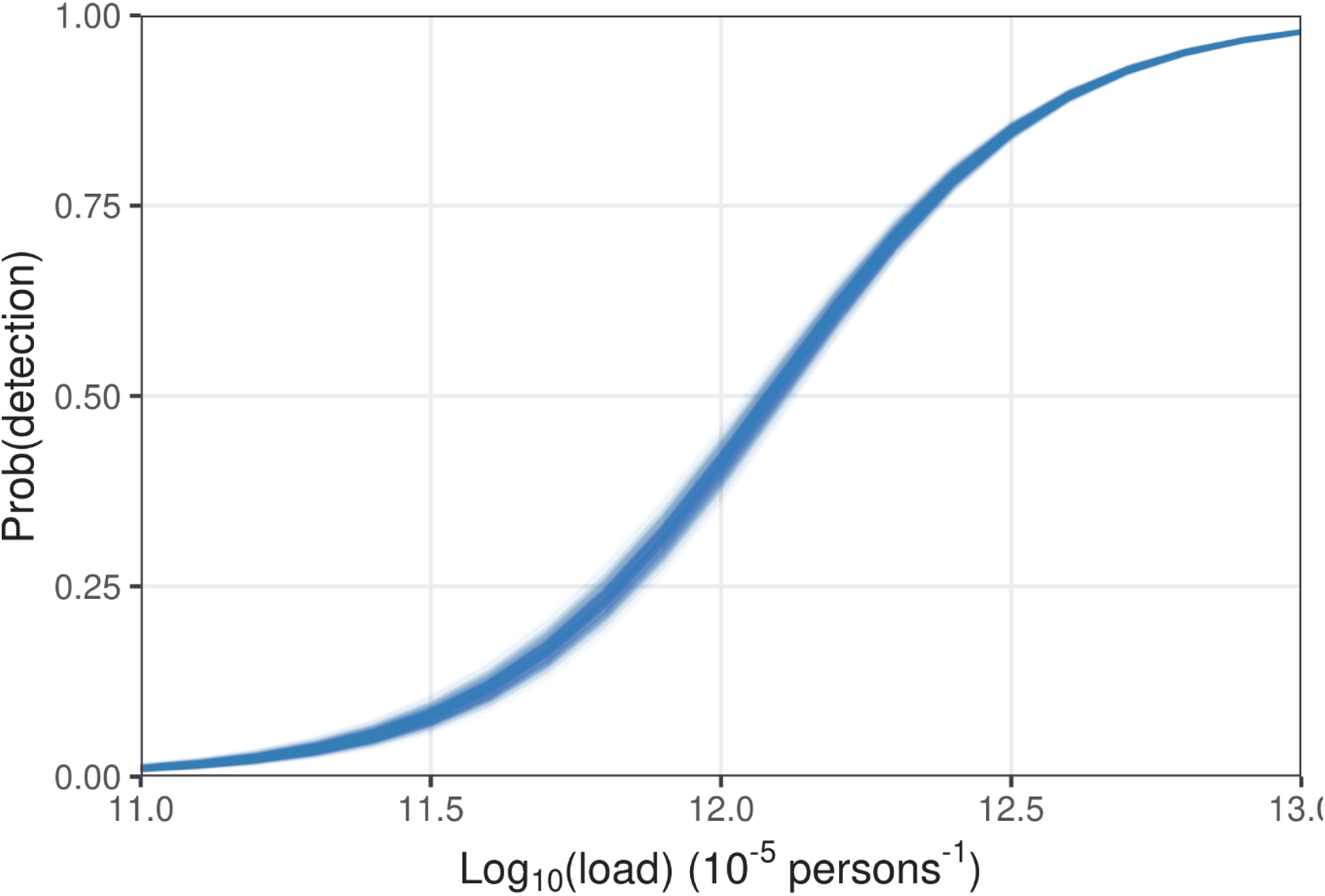
Estimated probability of SARS-CoV-2 RNA detection in sewage samples as a function of the (log_10_-transformed) sewage load. Shown are 1,000 samples from the posterior distribution.

From August 2020 until October-November 2020 virus loads increased by over an order of magnitude to a peak value close to approximately 10^14^ in many STPs. From this moment onwards virus loads started to decline gradually in all STPs. From May 2021 onwards, when the vaccination campaign was gathering strength, virus loads then started to decrease at a faster pace until the early summer. Subsequently, virus loads increased over the summer reaching a peak in early August 2021 by the temporary lifting of restrictions for vaccinated persons (colloquially called ‘Dancing with Johnson & Johnson’). Finally, from October 2021 virus loads started increasing again, first to just under 10^14^ in most STPs in December 2021, and subsequently to well over 10^14^ and even up to 10^15^ in February 2022.

Interestingly, the analyses can consistently uncover trends in virus loads that correspond to main epidemiological events. This is true even though measurement variation is estimated to be substantial. In fact, the posterior median of standard deviation of the observation model is *σ*=0.353 (95%CrI: 0.351-0.355, such that individual measurements can be 10^2*σ*^ ≈ 5.1 fold lower or higher than the estimated virus load. This, however, is still substantially smaller than the 100- to 1,000-fold exponential increases and decreases in virus loads from epidemic troughs to peaks and vice versa.

The estimated virus loads in the nine largest municipalities in the Netherlands are presented in Figure 4. These municipal estimates are obtained from the STP estimates by weighting the relevant posterior virus loads by the number of inhabitants in the focal municipality. The patterns are similar to those obtained in Figure 2 for the STPs. Again, we clearly observe the peaks and troughs at the municipality levels as estimated at the STP level. Estimated virus loads for all 345 municipalities are available in the online repository and are similar to the results presented in Figures 2 and 4.

**Figure 4.**
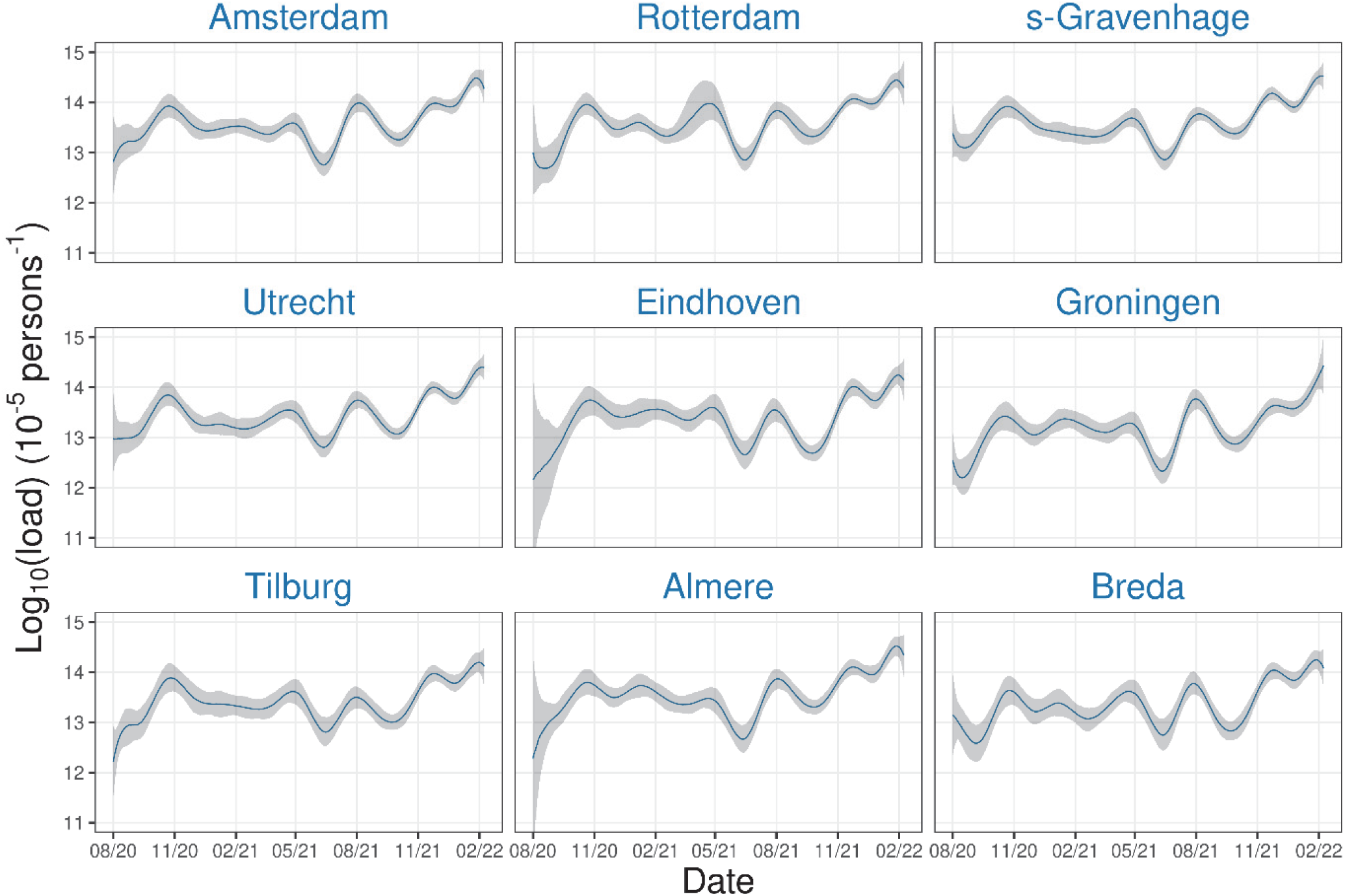
Estimated SARS-CoV-2 loads for the nine largest municipalities in the Netherlands. Municipalities are sorted by size (top left to bottom right). Total number of persons in these municipalities is 3,508948, representing 20.2% of the population (January 2020 census). Lines show the posterior medians of the virus loads with 95% credible intervals (grey bands). Full data for all 345 municipalities in the Netherlands are available at https://github.com/rivm-syso/SARS-CoV-2_sewage.

Finally, Figure 5 shows the inferred log-transformed virus loads in the Netherlands, using the population-weighted STP estimates, together with the log-transformed daily total number of hospitalisations (https://data.rivm.nl/covid-19/). Again, overall patterns of estimated virus loads are similar to those observed in Figures 2 and 4. Interestingly, epidemic troughs and peaks in the national estimate are less pronounced than those observed in most STPs. For instance, virus load estimates were as low as 10^12^ in many STPs in August 2020 but almost 10^13^ at the national level, and almost 10^15^ in the February 2022 peak but only just over 10^14^ at the national level. The observation that local epidemics are not fully synchronised is the reason that the national trend is less pronounced than those in the individual STPs (Figure 2). In fact, there is up to tenfold variation from one STP to the next in peak height and trough depth, and up to 2-4 week shifts in the timing of the peaks and troughs. Reassuringly, trends in estimated log-transformed virus loads correspond well with log-transformed daily hospitalisations. Interestingly, the number of hospitalisations at a given estimated virus load has decreased over time, and this decrease seems related to the rollout of the vaccination program in the first half of 2021. In-depth analysis of the combined SARS-CoV-2 STP and hospital data will be provided elsewhere (Hetebrij et al, in preparation).

**Figure 5.**
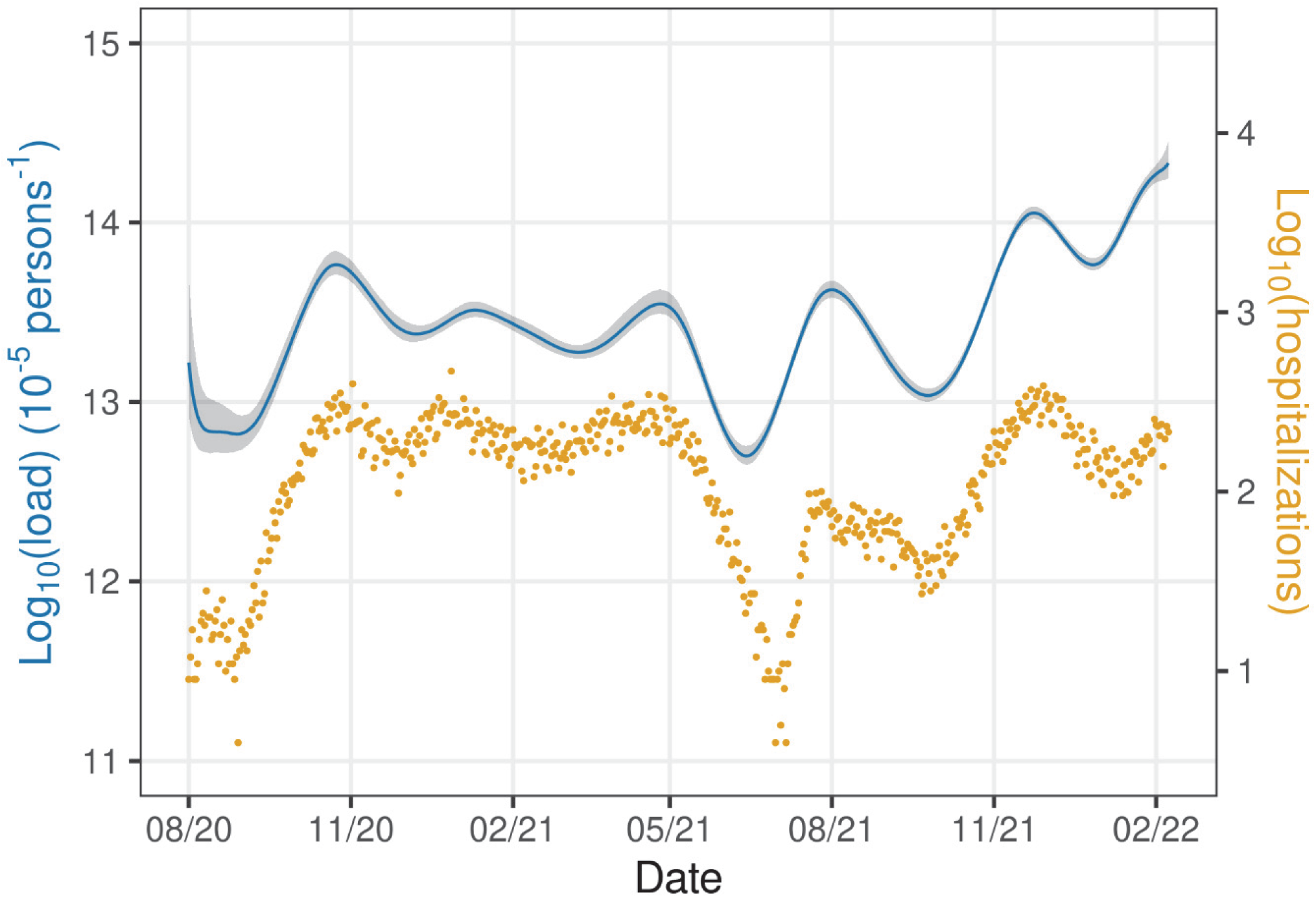
Estimated national log-transformed SARS-CoV-2 loads in sewage (line) with national log-transformed daily number of hospitalisations (dots). The line shows the posterior median of the STP-weighted national virus load with associated 95% credible band (shaded). Hospitalisations are retrieved from the open data provided by the National Institute for Public Health and the Environment (https://data.rivm.nl/covid-19/).

## DISCUSSION

Over 99% of the Dutch population is connected to the sewerage and thus to a STP. This makes it possible, in principle, to perform high resolution and near-real time surveillance of pathogens that are shed in feces, urine, and other excreta and secreta [8, 17]. However, sewage is a complex matrix to analyze [18], and proper controls are necessary to accurately quantify SARS-CoV-2 RNA. It is known that the amount of precipitation and the composition of industrial wastewater that is mixed in with the sewage of the general population will affect SARS-CoV-2 RNA concentrations determined by RT-PCR [19]. Still other factors that can affect measurements include the length of sewer lines and ambient temperature [20]. Here we have shown that even when there is substantial noise or day-to-day variation in RNA concentrations detected in sewage, it is possible to reliably estimate variations in underlying latent SARS-CoV-2 virus loads over time. This is possible because virus loads in sewage increase and decline exponentially during epidemic outbreaks, often by more than two orders of magnitude. These very strong variations over time dwarf the noise in the signal, which is determined by the standard deviation of the log_10_-transformed virus load measurements (*σ*). In the analyses, the standard deviation is estimated at *σ*=0.353, such that individual measurements can be approximately fivefold lower or higher than the estimated latent virus load.

Our analyses using multilevel splines provide a convenient research tool to analyse sewage data, as it enables a natural borrowing of information from the national virus load and samples taken around the same time. In this manner, we have been able to infer trends over time in the latent true virus loads for all 317 STPs in the Netherlands, as well as for a variety of other organisational levels (e.g., municipality, safety region, province, national). Specifically, the results provide estimates of the latent virus loads at all time points, and in particular for STPs on days that no measurements are available. In principle, our method of analysis can be extended further to any level of organisation that is built up from 4-digit postal codes in the Netherlands. In addition, subsampling from sewer lines in STPs with an unusually high virus load is also possible [21]. Especially in large STPs (such as Amsterdam-West and other STPs in Figure 2) there may be substantial differences between city districts with respect to SARS-CoV-2 prevalence, possibly associated with risk factors such as vaccination coverage and socio-economic status [22].

Although our analyses provide an adequate fit to the data, we do not claim that the results are in any way optimal for all SPTs at all time points. Rather, they provide a relatively parameter-sparse description of the data, linking the 317 STP time series through the national virus load while mildly favouring small STP-specific deviations from the national trend through the random effects. This seemed to work well in general, but it should be noted that the model has difficulty following very sudden changes in virus load data. A prominent example is presented by the sudden and strong drops in virus loads around October 2021 (Figure 2). This is only partially resolved by adding more knots to the splines, at the cost of greatly increasing computation times (not shown). Future extensions could also focus on adding STP-specific measurement noise, or perhaps even directly modelling the PCR data and water flow rates jointly. Here we would like to mention that it is not easy, even in the idealised context of large sample size, to select the optimal model from competing models using information criteria in a time series setting [23].

Our statistical modelling has provided a description of trends in virus loads at a local level by properly weighing all available wastewater measurements. In contrast, other modelling studies have focused on relating national or subnational virus loads to case notification data or hospitalisations using mechanistic modelling, with the aim to estimate the indicidence and prevalence of the number of infections over time ([24, 25] and references therein). The two approaches serve different purposes, and each has its strengths and limitations. A main strength of the mechanistic modelling is that all parameters have a biological interpretation, and that these analyses can be used for scenario studies. The results from such analyses, however, depend critically on model assumptions and are surrounded with large uncertainties. Our analyses do not yield a mechanistic interpretation but give precise estimates of latent virus loads that arguably are less dependent on specific model assumptions. In ongoing work, we aim to merge the two approaches by fitting transmission models at a local scale using generalized profiling [26].

Since the infrastructure of receiving sewage samples are in place, the detection of other viruses can be added to the Dutch sewage surveillance program. These might include rotavirus and enteroviruses but also influenza viruses [27, 28], thus providing a comprehensive surveillance tool for pandemic preparedness. Moreover, sewage surveillance for antimicrobial resistance has already shown its potential [29]. In principle, our methods of analysis can directly be applied to other targets and can deal with noise and unbalanced data in a principled manner.

The SARS-CoV-2 sewage surveillance program in the Netherlands has contributed to integrating available sewage data in a coherent framework, and also to informing the Dutch government on national and regional trends in SARS-COV-2 circulation [30]. Specifically, data at the national level have been used by the Dutch Ministry of Health, Welfare and Sport, and data from local analyses by the Municipal Health Services. At present, surveillance for SARS-CoV-2 is not only used for early detection of upticks in virus loads but increasingly also for SARS-CoV-2 surveillance in general, in view of increasing bias in testing data and decreasing numbers of hospitalisations. By integrating parameters such as age-stratified vaccination coverage and variant emergence, next aims are to explain age-specific hospitalisation admissions in the Netherlands from age-specific vaccination coverages, circulating variants, and virus load estimates in sewage (Hetebrij et al, in preparation).

## Data Availability

Data and code are publicly available at https://data.rivm.nl/covid-19/ (data) and https://github.com/rivm-syso/SARS-CoV-2_sewage (code).

https://github.com/rivm-syso/SARS-CoV-2_sewage

## ACKNOWLEDGEMENTS

We are indebted to representatives of the Dutch Water Authorities, in particular Mark van der Werf, Imke Leenen, and Bert Palsma, for facilitating the current study. We would like to thank all STP personnel working in water laboratories responsible for the actual sampling of wastewater. We gratefully acknowledge comments on the manuscript by Susan van den Hof and Rianne van Gageldonk-Lafeber. This research was financed by the Dutch Ministry of Health, Welfare and Sport, and by the Japan Society for the Promotion of Science (FM, JSPS KAKENHI Grant number 20J00793).

